# Mathematical Model Based COVID-19 Prediction in India and its Different States

**DOI:** 10.1101/2020.05.16.20104232

**Authors:** Jaspreet Singh, P. K. Ahluwalia, Ashok Kumar

## Abstract

By employing the successive approximation method to the real-time data of India and its different states, we have predicted the bounds of the spread of COVID-19 in India and its various states. The calculated lower and upper bound of patients (deaths) till 10^th^ June 2020 comes out to be 79496 (3835) and 241759 (7045), respectively. States like Delhi, Gujarat, Maharashtra, Punjab, Rajasthan and Tamil Nadu are the spike states as suggested by the range of expected COVID-19 patients and deaths. Impact of return of stranded pilgrims from Nanded (Maharashtra) has also been looked into in the case of Punjab. It has been found that Punjab may see ~ 5 times increase in the lower bound of expected patients till 10^th^ June 2020 due to the return of pilgrims from Maharashtra. Our study provides an insight into the possible number of expected patients and deaths in near future that may be of importance for the respective governments to be ready with the appropriate preventive measures and logistics to put appropriate infrastructure and medical facilities in place to manage the spread of deadly virus and go down the flattening curve.

## INTRODUCTION

The whole world is suffering from the COVID-19 pandemic which started from Wuhan, China [1]. So far due to this pandemic, the number of patients have risen to 4088848 and the death count risen to 283153 (mortality rate ~ 6.9%) [2]. The World Health Organization (WHO) declared COVID-19 a pandemic on 11 March 2020 by looking at its spread and threat to human life on the earth [3]. In India, the first case of COVID-19 was recorded on 30 January 2020 in Kerala, after that the number of cases rose to 69307 and death count rose to 2282 till 12^th^ May 2020. It has been confirmed that COVID-19 transmits from person to person through touching, coughing, sneezing [4], therefore, social distancing, isolation, and self-quarantine of suspected and infected patients are a must to avoid its spread. To control its transmission, lockdowns and curfews are implemented in various countries. In India the lockdown was implemented by the Government of India in the early stages of this pandemic in a country that restricted its fast exponential growth as has happened in China, USA, Italy, Spain, etc. [5-6]. However, the lockdowns and curfews are not the permanent solution of this pandemic and cannot be implemented for longer periods of time especially for a country like India, because of huge economic cost impacting vulnerable sections of the society, daily wage earners and large migrant labor. Beyond Lockdowns, every individual has to take his/her responsibility to maintain social distancing, use masks in public, avoid social gatherings, and avoid unnecessary movements to control its spread in the near future until its vaccine is discovered.

The main symptoms of this pandemic are high fever, dry cough, and body pain. It has been found recently that some cases are asymptotic [7] i.e. they do not show any symptoms. Such cases are very silent spreaders of the pandemic. It has been found that the virus can survive up to 3 hours in aerosols, 4 hours on copper, 24 hours on cardboard, and 72 hours on plastic and stainless steel [8]. This is the reason behind the exponential rise in cases of this pandemic worldwide despite the measures that have been taken to prevent its spread on war footing. The forecast of this pandemic based on the real-time data provides a way to help the governments and the concerned authorities to make the policies to minimize the infection of COVID-19 and halt it at the earliest with the participation of public at large. By anticipating the expected growth of patients, it is likely that governments provide required interventions such as COVID-19 health facilities, quarantine centers to screen suspected persons, dedicated COVID hospitals for infected cases and chalk out strategies to let the economic activities continue.

Mathematical models and data analytics are important tools to analyze and forecast the outbreak of infectious disease [9-11]. In this paper, we have utilized the successive approximation method [9] for short term forecast of infected and deceased people up to 10^th^ June 2020. We have calculated the lower and upper bound of patients and deaths till 10^th^ June 2020 for India and its various states. Our predictions hopefully can be helpful in social, economic, and health implications of COVID-19 for India at this stage. As India has a highly dense population and combining it with its Global Health Security Index (GHS) number (57/195) [12], India may be at great risk of this infectious disease.

## METHOD

Data were collected till 12^th^ May 2020 from the official website of COVID-19 (https://www.mygov.in/covid-19/) health advisory platform provided by the government of India. Data collected for the whole of India and its states, having patients count greater than 100 till 12^th^ May 2020 is analyzed with the help of a successive approximation method as employed in reference [9, 11]. We find the spread (death) ratio as the ratio of the number of cases (deaths) on a particular day to the sum of the cases (deaths) on that day and the next day. We use these ratios to find the future number of patients as well as deaths till 10^th^ June 2020.

Suppose *a_i_* are the number of cases at day *i* than we define the ratio as:

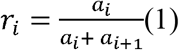

For the next day *i +* 1, the ratio is given by:

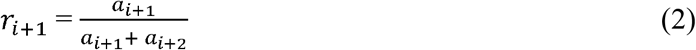

We use these ratios to find the future state of cases as:

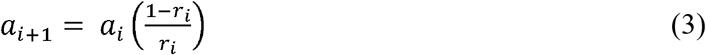

From eq. (3), it is clear that if *r_i_* → 0, then there is a large increase in the number of new cases and if *r_i_* →1, the number of new cases significantly decreases. Using this technique, we have found the spread ratio and death ratio by which we have predicted the number of patients and deaths till 10^th^ June 2020. The lower bound of cases is found by selecting the maximum *r_i_* i.e.

For lower bound of cases, we use *r_i_* as:

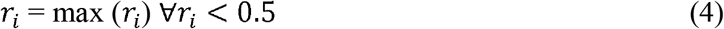

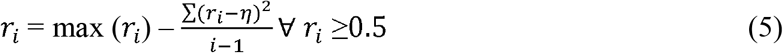

where *η* is the mean ratio given by:

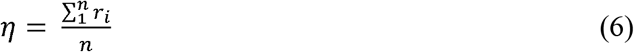

By using this technique, we have forecast the spreading of COVID-19 in India and its different states.

## RESULTS AND DISCUSSION

We have plotted the spread and death ratios w. r. t number of days in Figure 1a and the number of expected patients and deaths till 10^th^ June 2020 in Figures 1b and 1c, respectively. The spread and death ratios vary nearly around 0.475 that indicates the spread of this pandemic in India. The calculated lower and upper bound of patients (deaths) till 10^th^ June 2020 comes out to be 79496 (3835) and 241759 (7045), respectively. It is clear that if the prevention measures are strictly followed, then there are reasonable chances of controlling the spread of this pandemic in the country, otherwise, the number of patients and deaths may rise exponentially in a no time. We have also done the same calculations for different states of India. The lower and upper bound of patients and deaths are listed in Table 1. We have plotted the pandemic trends for different states of India in Appendix. The states like Delhi, Gujarat, Maharashtra, Punjab, Rajasthan and Tamil Nadu are at much greater risk as suggested by the range of expected patients and deaths (Table 1).

**Figure 1.**
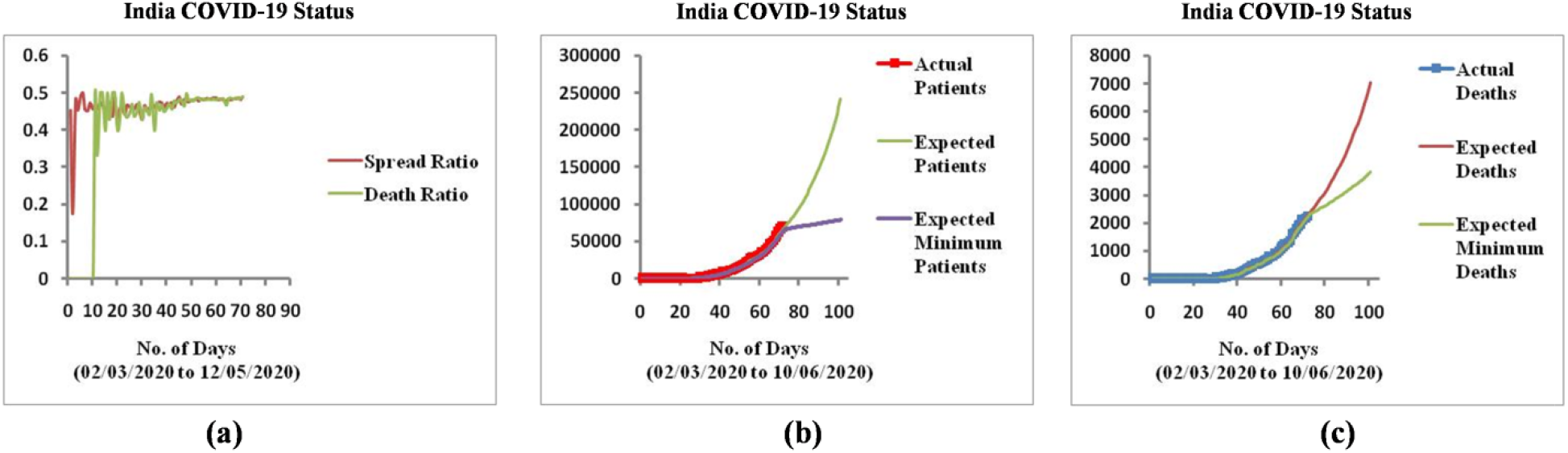
**(a)** Spread and death ratio of COVID-19 in India. **(b)** Prediction curves for expected patients of COVID-19 in India. (c) Prediction curves for deaths of COVID-19 in India.

**Table 1:** Number of expected patients and deaths due to COVID-19 in different states of India till 10^th^ June 2020.

| S.No. | State | Expected Patients (Range) | Expected Deaths (Range) |
| --- | --- | --- | --- |
| 1 | Andhra Pradesh | 2549-3467 | 109-349 |
| 2 | Bihar | 870-5853 | 7-63 |
| 3 | Delhi | 8312-25829 | 133-571 |
| 4 | Gujarat | 9261-26876 | 991-1625 |
| 5 | Haryana | 760-2097 | 28-174 |
| 6 | J&K | 1037-1544 | 21-212 |
| 7 | Jharkhand | 210-275 | 8-45 |
| 8 | Karnataka | 898-1360 | 53-80 |
| 9 | Kerala | 591-766 | 8-33 |
| 10 | Madhya Pradesh | 4376-14092 | 467-491 |
| 11 | Maharashtra | 28173-109372 | 1590-2965 |
| 12 | Odisha | 614-6327 | 7-31 |
| 13 | Punjab | 2548-48048 | 60-214 |
| 14 | Rajasthan | 4513-14039 | 320-550 |
| 15 | Tamil Nadu | 10467-166966 | 103-1728 |
| 16 | Telangana | 1556-8309 | 73-80 |
| 17 | Uttar Pradesh | 3703-8132 | 172-767 |
| 18 | West Bengal | 2132-12514 | 440-833 |

It is pertinent to note that in Punjab, the number of cases up to 1^st^ May 2020 were 357 and within one week from this the cases rose to ~ 1700. This sudden rise in the number of positive cases was due large number of pilgrims (~3500) returning to Punjab from Nanded Hazur Sahib, Maharashtra [13]. We have predicted the pandemic status of Punjab from two sets of data up to 1^st^ May and 12^th^ May, to look into the effect of Nanded pilgrims in the expected number of cases (Figure 2a, 2b). From the data up to 1^st^ May 2020, the expected lower and upper bound of patients (deaths) till 10^th^ June 2020 are 527 (58) and 26115 (165), respectively, whereas including data up to 12^th^ May 2020, the lower and upper bound of patients (deaths) rises to 2548 (60) and 48048 (214), respectively, which predicts a significant increase in the number of cases (~5 time increase in the lower bound and ~2 time increase in the upper bound of number of expected patients up to 10^th^ June 2020). Moreover, the mortality rate is higher in Punjab (19 deaths out of 357 patients (~18%) as on 1^st^ May 2020) as compared to ~3.5 % of country. This may make the situation alarming if the spread of pandemic is not controlled. Similar things also happened in other states. The gathering of thousands of migrant workers at the bus terminal of Anand Vihar of Delhi, similar gathering at Bandra station in Mumbai, Tablighi Jamaat event made the situation more difficult to control.

**Figure 2:**
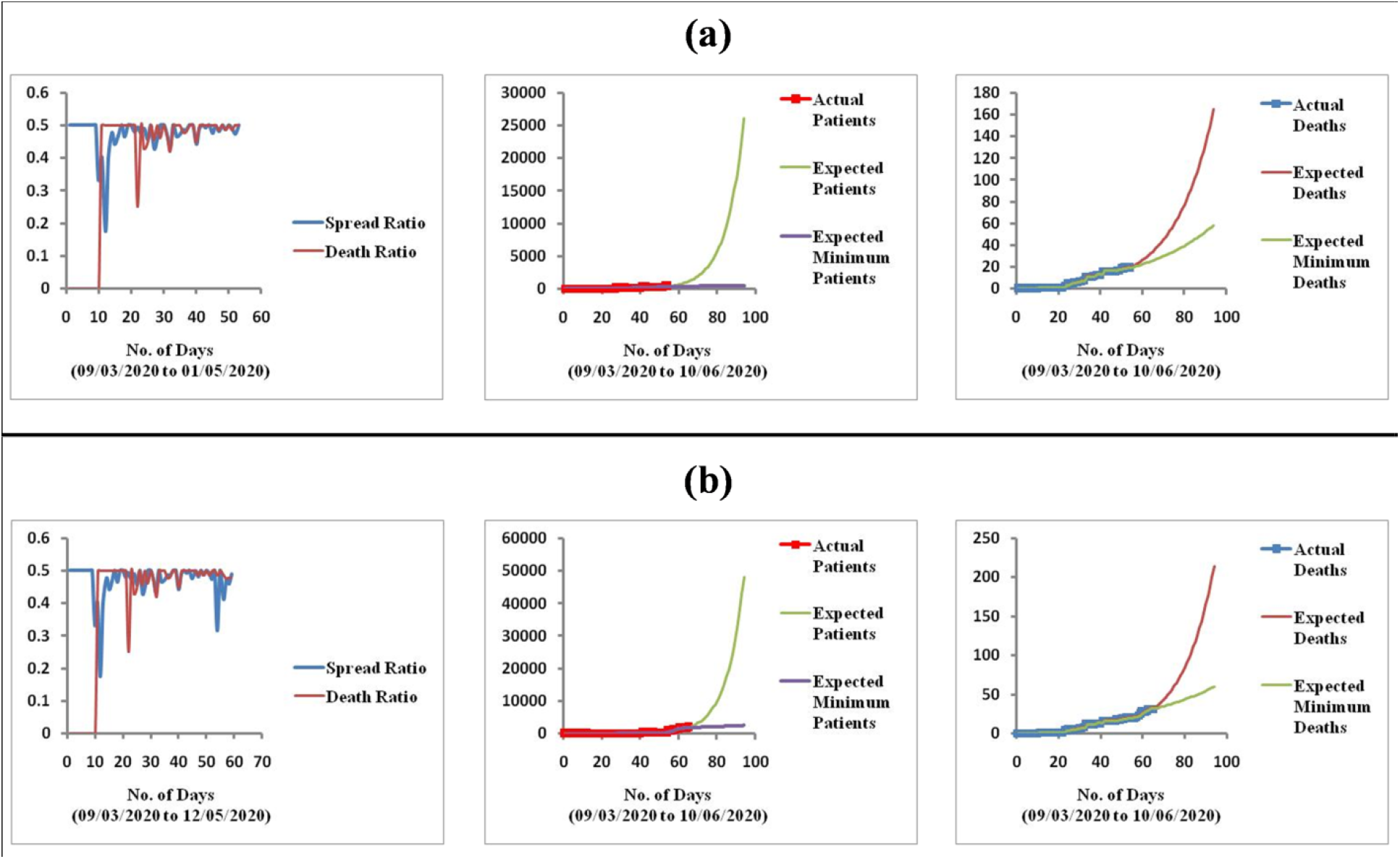
Prediction of COVID-19 status of Punjab with data**(a)**up to 1^st^ May 2020**(b)**up to 12^th^ May 2020.

The mathematical model that we have used is based on real-time data and that data is collected when the country has already taken preventive measures such as lockdowns and curfews. Proper social distancing and quarantine are the only successful means of controlling the spread of this pandemic. The rapid and large-scale testing of suspected cases is a must to prevent its spread. Isolation of suspected cases, social distancing and quarantine should also be strictly followed in India to prevent the spread of this pandemic. Our results may provide insight into the course of spread of COVID-19 in India and its different states, and may be of importance to central government and governments in the states and to take appropriate preventive measures to stop the spread of the COVID-19 till the vaccine is discovered, a scenario which is unlikely to be in immediate future.

## CONCLUSIONS

In summary, India maybe at high risk due to its highly dense population and moderate health facilities (ranks 57/195 in Global Health Security Index). Though we have done well compared to many countries till date, but the future is full of uncertainties. We need to draw appropriate lessons from the predictive nature of studies as has been carried out in this paper with clear indications of the likely spread. Also, this study clearly indicates the likely hotspot states where immediate interventions will be needed to break the COVID-19 spread. Our results we hope will help the governments to plan their strategies against the spread of COVID-19. There is lot of implication of increase in testing facilities on the forecast made in this paper

## Data Availability

N/A

## Appendix

The plots of Spread ratio, death ratio, expected patients and deaths in different states of India.

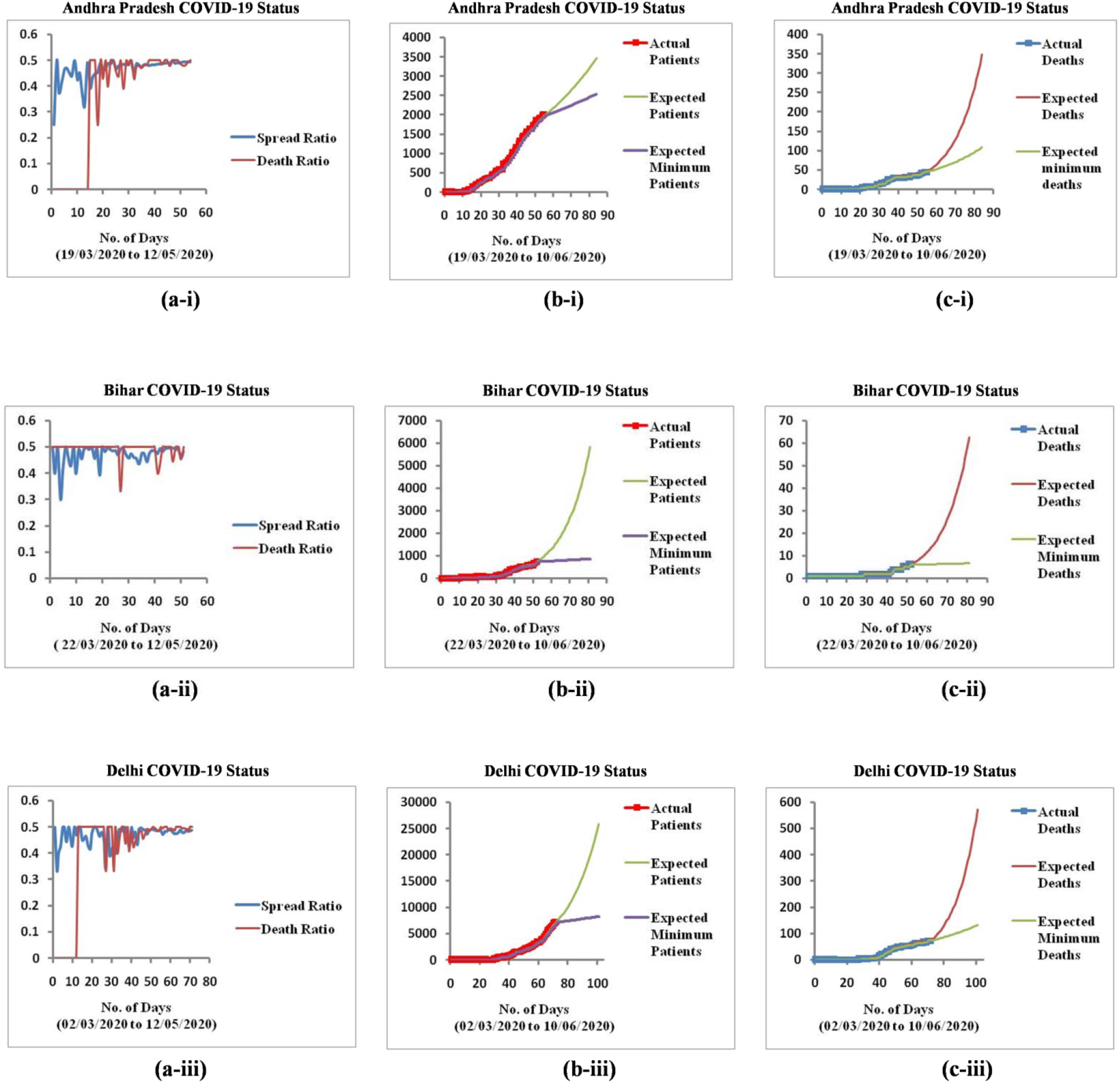

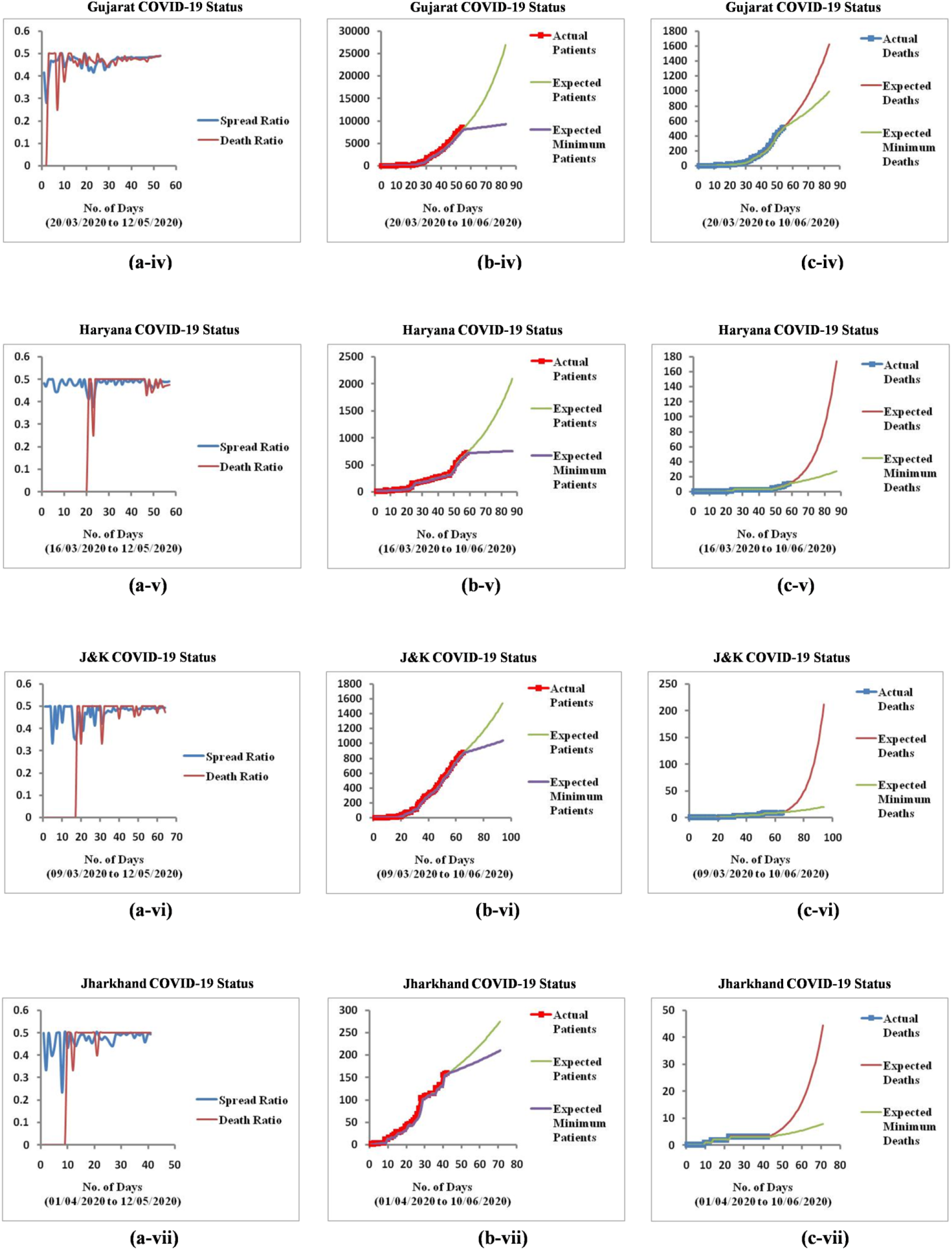

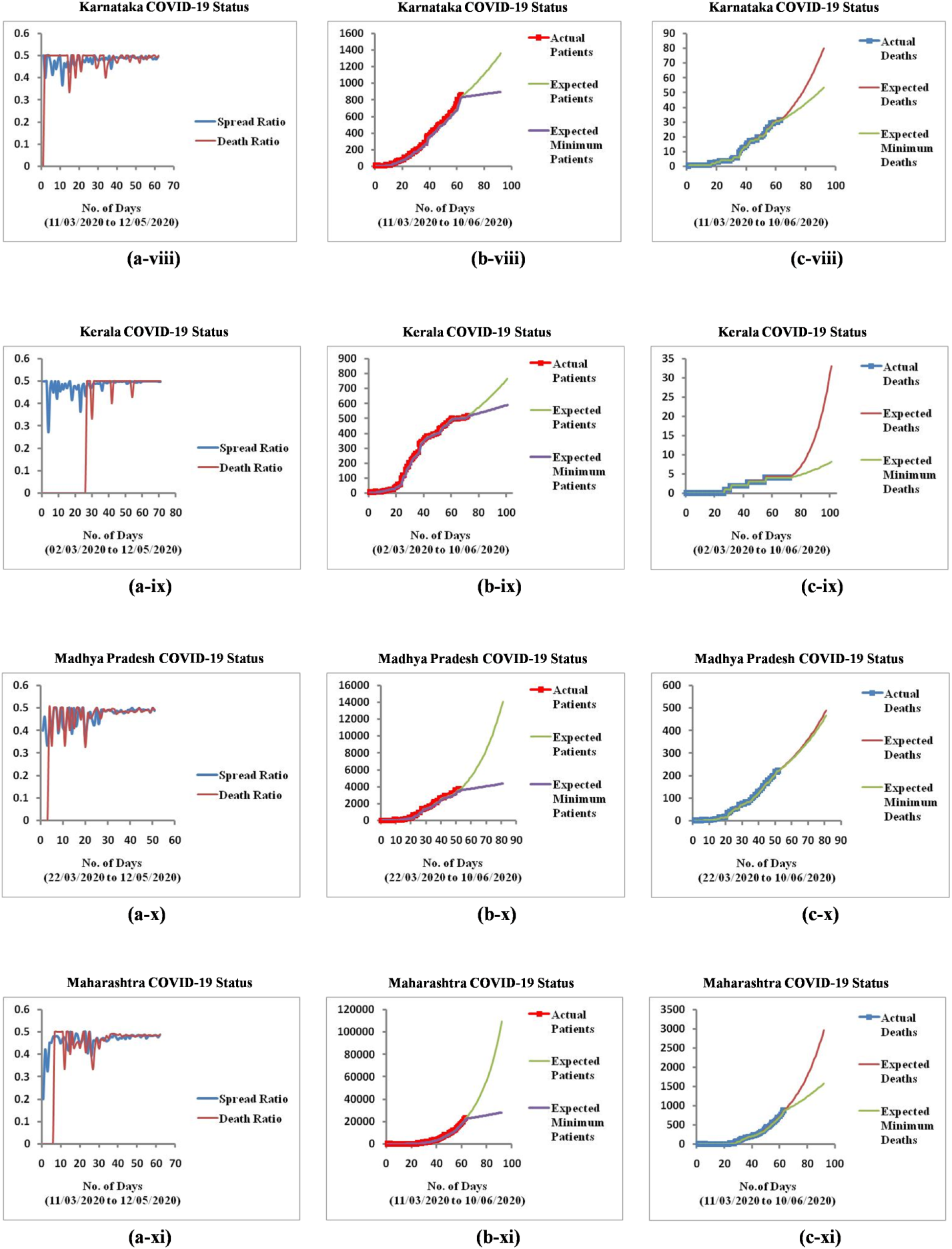

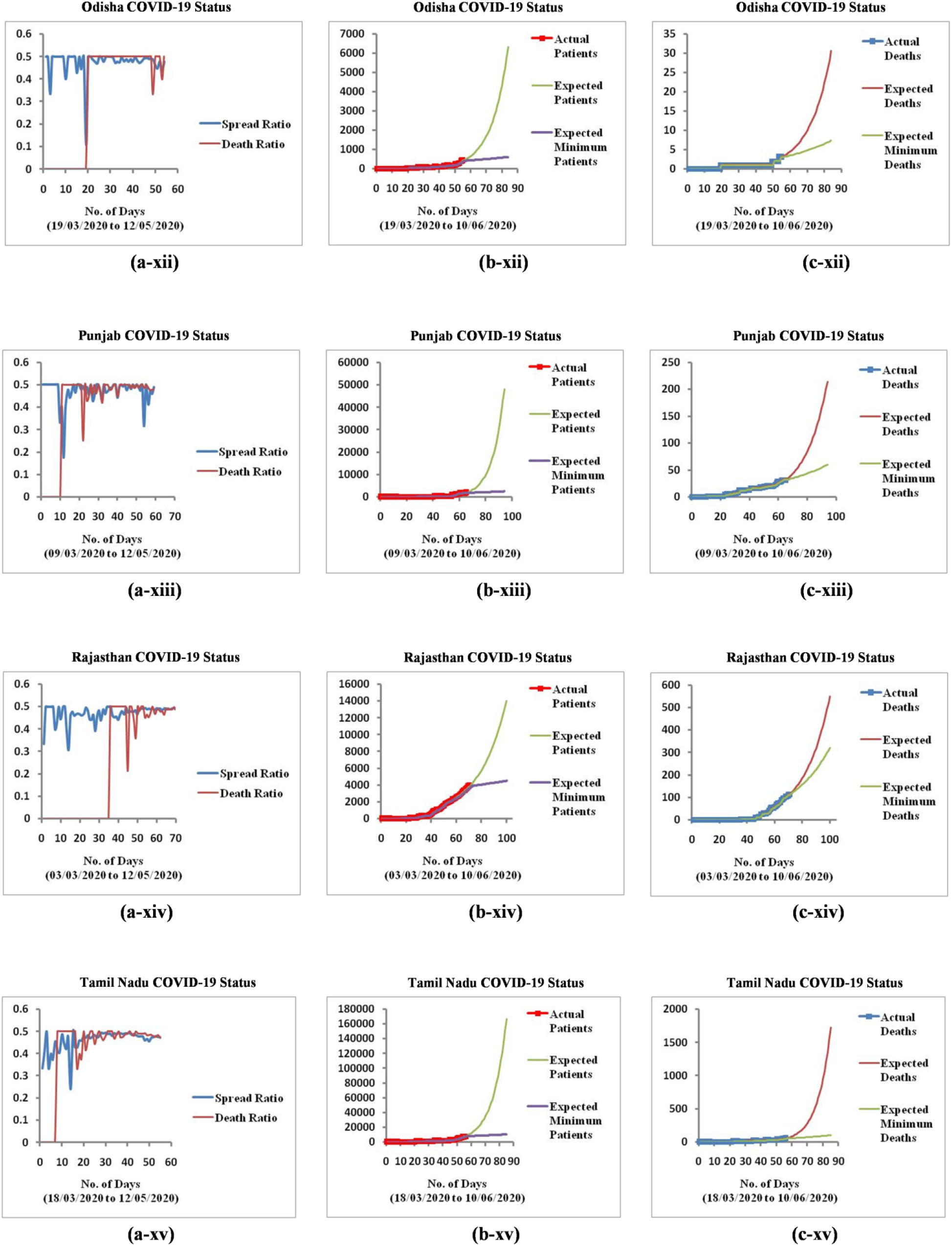

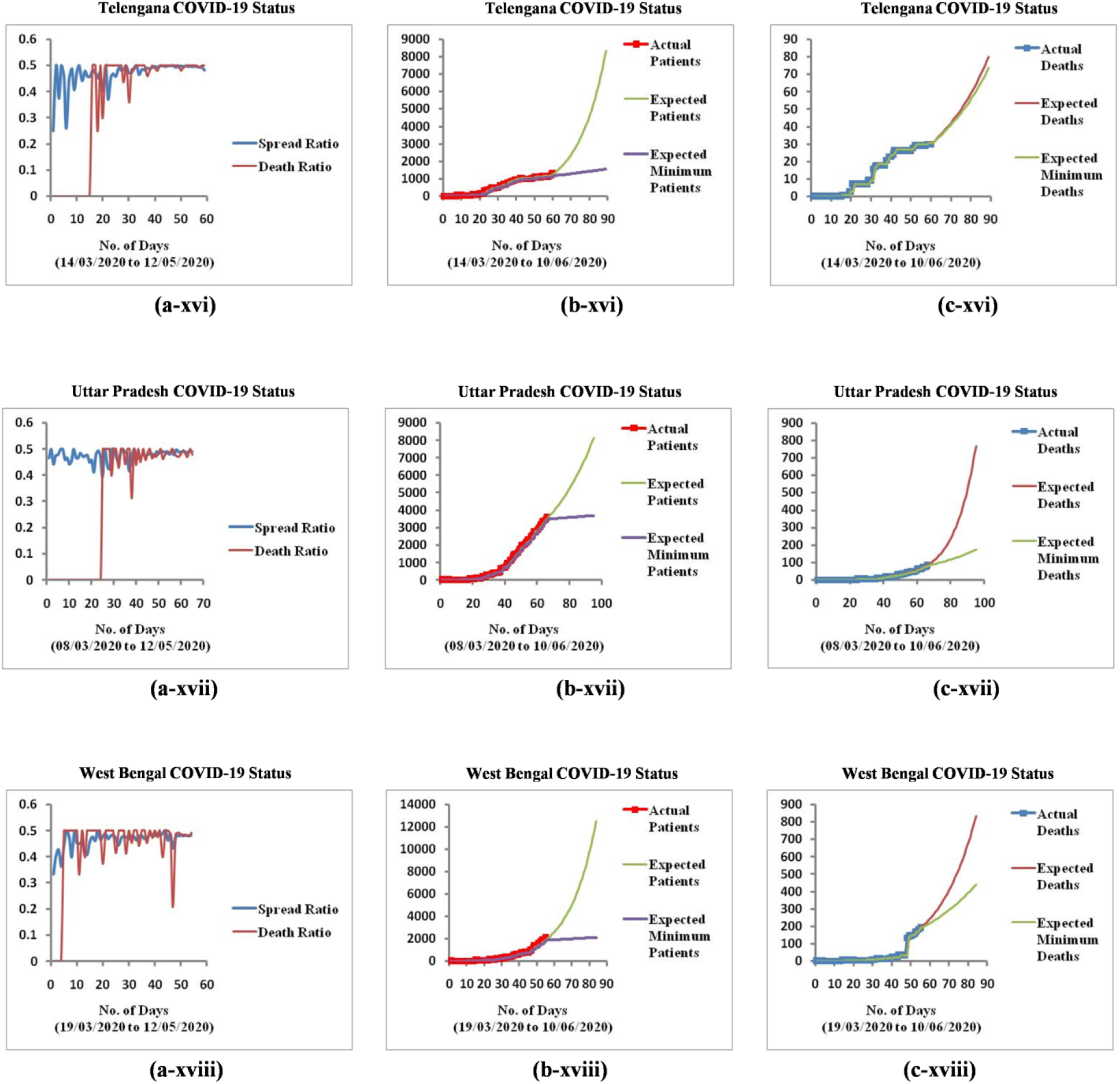

